# VaaS is a Multi-Layer Hallucination Reduction Pipeline for AI-Assisted Science: Production Validation and Prospective Benchmarking

**DOI:** 10.64898/2026.03.24.26348935

**Authors:** Ankit Sabharwal, Milit S. Patel, Anna Carrano, Maarten Rotman, Wesley Wierson, Stephen C. Ekker

## Abstract

The deployment of large language models (LLMs) for science carries an intrinsic risk: hallucination of citations, fabricated drug approvals or clinical trials, and unsupported experimental outcomes. Here we describe the testing and deployment of a novel systematic, multi-layer approach called the *Validation as a System* (VaaS) pipeline, iteratively developed during the construction of an open-source, living Rare Disease Database (RDD). We report lessons learned and production results from 225 carefully annotated rare disease gene curations and a prospective 100-gene collection (99 net new), together representing over 3,000 verified citations. After three iterations of directed refinement, the net functional hallucination rate approached zero. We validated the pipeline using three complementary benchmarks: (1) VaaS-RIKER2, a 640-run prospective ablation study (4 conditions × 4 temperatures × 40 genes) plus 117 open-weight model runs on dedicated GPU hardware — unguided LLM output produced 95.9% Type II hallucination (wrong-topic citations that exist as real papers but carry a correct claim context yet do not support the cited claim); the full VaaS protocol achieved 0.0% Type I and 6.5% Type II, a *>*14-fold reduction; live PMID verification alone (C3) eliminated both error types entirely (0.0%/0.0%); (2) an independent L3 citation audit of Wave 3 (179 PMIDs, 99.4% valid, 0 Type I errors); and (3) the MedHallu clinical hallucination benchmark, on which the VaaS protocol achieved F1 = 0.9853 on the hard tier (cases where all benchmark ensemble models were fooled), compared to the published GPT-4o baseline of F1 = 0.811 (Pandit et al., 2025). Three independent open-weight models (`llama3.2`, `qwen2.5:14b`, `mistral:7b`) showed 81–87% Type II rates under unguided conditions, confirming that wrong-topic citation hallucination is structural and model-agnostic. In contrast, the corresponding VaaS rate was measured to be zero (*n* = 508 verified citations; 160 runs, C4 full protocol) under the same conditions. Human validation of ≥ 50 entries confirmed zero Type I errors and less than 0.5% Type II errors in the manual curation test. The VaaS pipeline operated at less than ∼$1 overall per comprehensive gene review, demonstrating that citation-integrity standards in AI-assisted biomedical synthesis are achievable at production scale. The VaaS approach represents, to the authors’ knowledge, the lowest measured hallucination system for science to date and is set to further accelerate the use of AI and AI agents for advancing research.

## 1 Introduction

Large language models (LLMs — AI systems that generate text by predicting the next token in a sequence, trained on large corpora of scientific and general text) have demonstrated strong capability for scientific text generation, including literature synthesis, grant writing, and disease review. However, LLMs are prone to a class of errors particularly damaging in science: confident fabrication of citations, incorrect drug approval statuses, fake clinical trials, and fictitious experimental outcomes. Unlike errors in casual text, a false PMID or a fabricated FDA approval annotation in a scientific product could mislead real research decisions — influencing grant hypotheses, clinical guidance, and competitive landscape analyses.

Hallucination in LLMs is not a correctable bug but an architectural property of next-token prediction training, a core technology that as a feature rewards confident output generation over calibrated uncertainty. As OpenAI has noted in its technical documentation and as independently confirmed in peer-reviewed studies of LLM citation accuracy, next-token prediction objectives embed hallucination as an optimization incentive (OpenAI et al., 2023; Ji et al., 2023). Longitudinal benchmarks have confirmed the persistence of the problem: initially up to 47% of ChatGPT-generated medical references were entirely fabricated (Bhattacharyya et al., 2023), and GPTZero detected 100+ hallucinated citations (Type I errors — citations that do not exist in any publication database) in accepted NeurIPS 2025 papers (Goldman, 2026). For many, these errors are considered an acceptable trade-off when working with LLMs.

Type II citation hallucinations — defined as wrong-topic citations that exist as real papers but carry a correct claim context yet do not support the cited claim — share characteristics with structural hallucination as described by Boudourides (2026), in which relational structure rather than surface content is distorted.

The depth of this technical limitation of hallucinations in even the most advanced LLMs such as Claude Opus 3.5 was immediately apparent upon the launch and deployment of new, open-source agentic systems (data not shown), with full reports provided with confidence by the AI agents but filled with nonsensical research findings not grounded in any facts. To address this problem, UViiVe used a novel deployment of OpenClaw to develop a fleet of AI agents with unique skills to tackle these intrinsic hallucinations after weeks of internal training. This AI fleet was subsequently deployed in partnership with the Center for Rare Disease at Dell Medical School in February–March 2026 to build a systematic rare disease gene review and knowledge database: initial 225 entries covering mitochondrial, neurological, lysosomal, cardiac, sensory, immune, and renal disease genes, produced across three further iterations. The first 225 used the baseline advanced agentic system that predated the full Aletheia epistemic integrity protocol; the next 100 genes were subsequently generated post-Aletheia (99 net new entries; one gene was re-reviewed as an internal quality check against Wave 1/2 output) and was used to prospectively challenge this revised approach. Each entry required accurate PMIDs, correct clinical trial status, and honest representation of therapeutic approval status. At this scale (∼3,000 total citations), even a 3% fabrication rate would embed approximately 90 false references into a product intended for grant writing, lab meeting preparation, research development and NotebookLM-assisted querying. The goal was to achieve as close to a net functional hallucination rate of zero.

The VaaS (Validation as a System) pipeline is an agentic-led, multi-step automated workflow in which AI agents execute sequential or parallel subtasks with defined quality gates, and addresses citation hallucination through layered verification at the point of claim commitment. This paper describes the iterative quality improvement process across three production phases, with quantitative measurements at each stage. To prospectively validate the pipeline, we conducted three complementary benchmark studies: (1) a controlled stress test battery (Study ID VaaS-HT-001; five genes under both unguided and protocol conditions, plus three challenge tests); (2) the VaaS-RIKER2 prospective benchmark — a 40-gene × 4-condition × 4-temperature study generating 640 controlled runs plus an open-weight model arm comprising 117 runs across three independent model architectures; and (3) the MedHallu clinical hallucination benchmark (Pandit et al., 2025); results are reported in Section 3.7. Note: the term “VaaS-RIKER2” refers to validation of the VaaS pipeline using the RIKER benchmark framework (Roig, 2026) (arXiv:2603.08274); RIKER is an independent external benchmark, not a component of the VaaS architecture.

### Related work

The Machine Learning (ML) community has developed several families of hallucination mitigation techniques. Retrieval-Augmented Generation (RAG) — the dominant paradigm for grounding LLM outputs in external verified data — retrieves relevant documents from databases before generation, reducing hallucinations substantially in specialized deployments. In a controlled evaluation of RAG-augmented cancer-information chatbots, hallucination rates dropped from ∼ 40% without RAG to 0% (GPT-4) and 6% (GPT-3.5) when grounded in a curated knowledge base (Nishisako et al., 2025). CiteGuard (Gangwani and Bansal, 2025) is the closest published analog to our PMID verification approach, achieving citation-level F1 = 0.89 on a 7,221-citation benchmark. VeriCite (Qian et al., 2025) uses a three-stage natural language inference (NLI) pipeline for RAG citation quality improvement. Multi-agent systems reduce errors by 0–42% compared to single-agent architectures in controlled studies; this figure, derived from a radiology-specific study (Salehi et al., 2025), is used here as a general reference point for multi-agent validation effects — direct applicability to literature mining pipelines may vary.

The RIKER2 benchmark (Roig, 2026) provides a complementary characterization of open-weight model hallucination across multiple task types and context lengths. Their best-performing open-weight models achieved 0% hallucination rate on the grounded reading task (answer present in document); their ungrounded citation generation task, which mirrors our C1 (unguided baseline) condition, showed substantially higher rates. We situate our findings against RIKER2 baselines in Section 3.10.

The MedHallu benchmark (Pandit et al., 2025) provides an orthogonal test of hallucination detection in medical question-answering. Comprising 10,000 PubMedQA-derived question–answer pairs stratified into easy, medium, and hard tiers, its hard tier is operationally defined as cases where all LLMs in the evaluation ensemble are fooled — representing the most clinically consequential failure mode. The best published model (GPT-4o with knowledge augmentation) achieves F1 = 0.811 on the hard tier. We evaluated the VaaS protocol against this benchmark including using the latest LLM models such as Claude Opus 4.6; results are reported in Section 3.7.

### What is novel

Against this backdrop, the specific contributions of the VaaS pipeline are: (1) production deployment at biomedical scale — 225 advanced disease gene analyses at estimates of less than $1 in API costs per detailed assessment; (2) an epistemic integrity constraint embedded in the agent’s system prompt (described further in Section 2.1); (3) a self-improving corrections list injected verbatim into every agent briefing; (4) drug approval hallucination as a named, characterized error category; (5) tiered risk assignment stratifying entries by hallucination risk; (6) a prospectively designed benchmark (VaaS-RIKER2) providing the first controlled 4-condition ablation study of a production hallucination-reduction pipeline in rare disease AI synthesis; (7) open-weight model characterization on dedicated local GPU hardware; and (8) hallucination topology characterization by gene tier. We situate this work within the current literature and reflect on what remains unsolved from our vantage point.

## 2 Methods

### Overview

All of this work was initially established by the authors and then deployed in collaboration with a series of advanced and custom-trained AI agents that then served as legitimate co-contributors. The main charge throughout was to develop an approach that handled the inherent failed integrity layer when core facts would change by an LLM. The resulting method described herein is a hybrid research product and process with input by both human and AI scientists.

### 2.1 Epistemic Integrity Constraint (“The First Law”)

The pipeline incorporates an epistemic integrity constraint (colloquially, “The First Law”) embedded in the agent’s system prompt, which prohibits generation of unverified citations. This is an operationalization of standard RAG practice rather than a novel methodological contribution. The constraint reads, in relevant part:

> *Never fabricate data, results, citations, or conclusions. Do NOT invent PMIDs, DOIs, or citations. Do NOT fabricate statistical results, p-values, or effect sizes. Do NOT hallucinate experimental outcomes. When uncertain, quantify uncertainty*.

Its placement at the identity level (rather than as a task-level instruction) was deliberate: it frames scientific honesty as a core attribute of the agent, not a rule that competing instructions might override.

**Figure 1.**
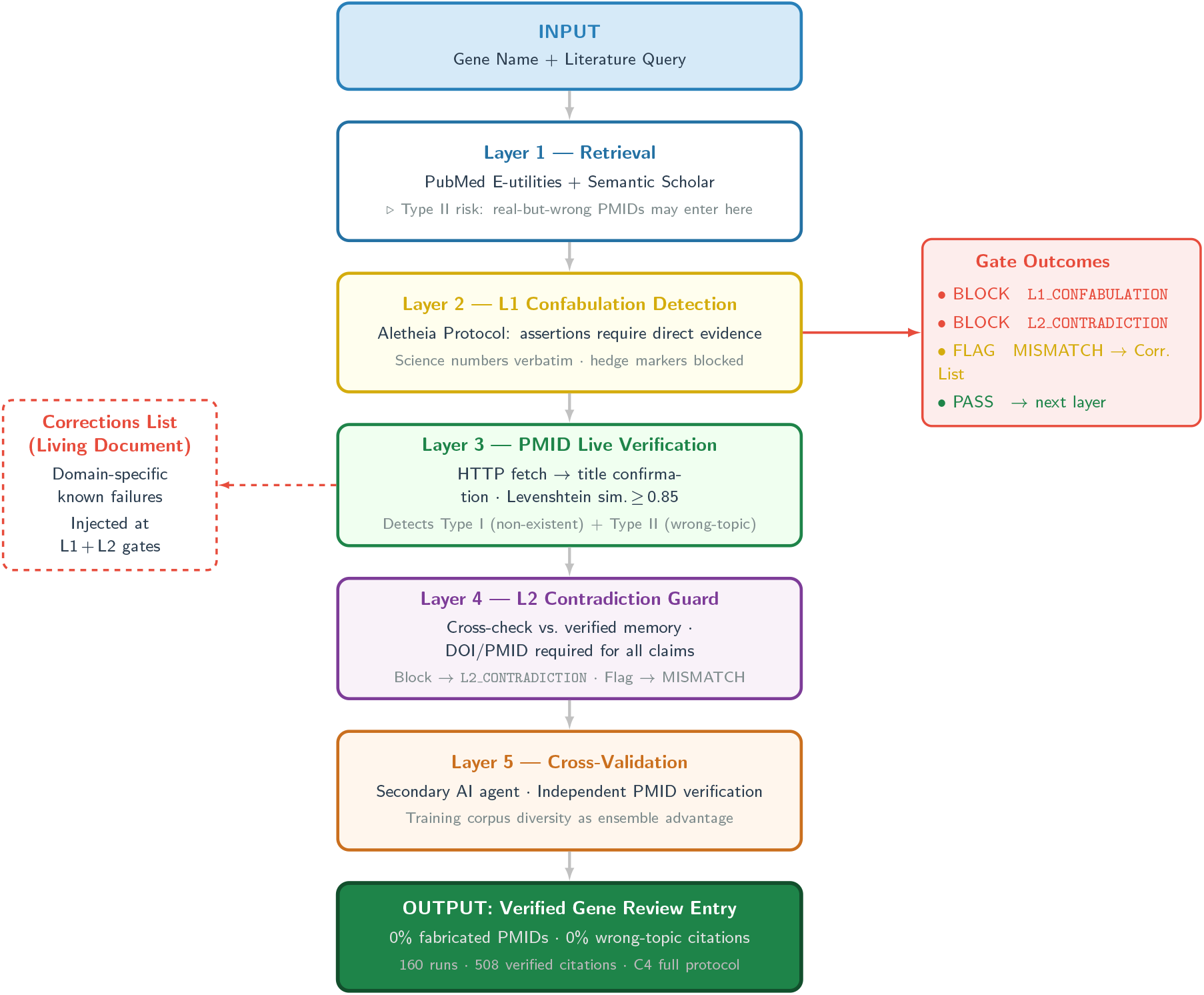
VaaS multi-layer pipeline architecture. Schematic of the sequential verification stages from raw LLM output through epistemic integrity constraint, live PMID fetch, topic verification, and corrections-list injection to final curated output.

### 2.2 PMID Verification Protocol

All disease review subagents were instructed to verify every cited PMID by direct HTTP fetch: GET https://pubmed.ncbi.nlm.nih.gov/[PMID]/

- **Parse:** title, first author, journal, year
- **Confirm:** matches the claim being cited
- **Flag:** `[WARNING] SECONDARY SOURCE` if only cited from a review
- **Reject:** `[REJECT] REJECTED` if fetched paper is clearly wrong topic

This protocol required the agent to make a verifiable claim (live fetch + title match) rather than rely on parametric memory. The distinction is critical: parametric recall of a PMID is a hallucination risk; a live HTTP fetch with title confirmation is not.

In the event of rate limiting or bot-blocking by PubMed or other publishers, the pipeline implements exponential backoff with a maximum of 3 retries. Persistent access failures are logged and flagged for manual review, with affected citations marked as `unverified-access` rather than silently excluded.

### 2.3 Formal Citation Error Taxonomy and Benchmark Prompt

#### Type I error — Citation Fabrication

A cited PMID does not exist in PubMed. This is detectable only by live fetch. Type I errors represent complete confabulation of a citation identifier.

#### Type II error — Citation Hallucination

A cited PMID exists and resolves to a real paper, but that paper does not support the claim being cited (wrong topic, wrong gene, wrong finding). Type II errors require topic-gene consistency checking beyond simple existence verification. Our Type II error category shares characteristics with structural hallucination as described by Boudourides (2026), in which relational structure rather than surface content is distorted.

These error types are operationally distinct and are counted independently throughout this manuscript. The VaaS-RIKER2 benchmark provides the first prospective ablation study of interventions targeting both error types independently.

#### Controlled benchmark prompt

The exact prompt used as the standardized benchmark input across all production gene reviews is:

> “Generate a comprehensive rare disease gene review for `[GENE]`, including OMIM disease number, inheritance pattern, clinical features, zebrafish model relevance, and 3 primary literature citations with PMIDs. Verify each PMID by direct HTTP fetch before including it. Apply the epistemic integrity constraint at all times: do not fabricate any citation, PMID, drug approval status, or clinical trial outcome.”

Each gene review targets a minimum of 2 and maximum of 5 primary literature citations, prioritized by recency (≤5 years where available), citation impact, and direct relevance to the disease mechanism described.

### 2.4 Topic Verification Protocol

To address the distinction between existence errors (Type I) and topic-relevance errors (Type II), the VaaS pipeline incorporates a dedicated topic verification step. After a PMID is confirmed to exist via live fetch, the fetched abstract is evaluated against the target gene using the following standardized prompt:

#### Topic Verification Prompt (verbatim)

> “Given the following paper abstract, does this paper directly study `[GENE_SYMBOL]`? Answer YES, NO, or BORDERLINE with one sentence of explanation. Abstract: `[ABSTRACT]`”

This prompt is applied to every PMID candidate before inclusion in a gene review. The three-tier response maps directly onto the citation disposition system:

- **YES** → citation included as a verified hit
- **NO** → citation rejected (wrong-topic category)
- **BORDERLINE** → citation flagged for human review; included with a `[WARNING]` qualifier and noted in the audit log

#### Worked examples

- **Example 1 — Clean Hit (YES):** Gene: *POLG*. Abstract: “We report three novel pathogenic variants in the *POLG* gene in patients presenting with mitochondrial DNA depletion syndrome… “ Response: YES — This paper directly studies *POLG* gene variants and their pathogenic consequences.
- **Example 2 — Wrong-Topic Mismatch (NO):** Gene: *POLG*. Abstract: “Mitochondrial disease encompasses a heterogeneous group of disorders affecting the respiratory chain… “ Response: NO — This paper covers mitochondrial disease broadly and does not specifically study the *POLG* gene.
- **Example 3 — Borderline (BORDERLINE):** Gene: *POLG*. Abstract: “Pathogenic variants in mitochondrial polymerase pathway components produce variable neurological phenotypes… “ Response: BORDERLINE — This paper studies the *POLG*-associated pathway but does not directly name or characterize the *POLG* gene itself.

### 2.5 Self-Improving Corrections Injection

As errors were caught across production, a known-corrections list was compiled and injected into every new subagent briefing. The list evolved from 0 items (Pilot) to 14 items (Wave 2) to 20+ items by the end of production. The corrections list applied in VaaS-RIKER2 (C2 and C4 conditions) included: Valproate contraindication in POLG-related disease; Cerliponase alfa (Brineura) approved for CLN2 only (not CLN3); GARS1 autosomal dominant inheritance; Lumevoq EMA approval withdrawn April 2023; Setrusumab Phase 3 failure December 2025; ADCK3 = COQ8A (synonym); BCS1L2 does not exist as a gene; CoQ10 off-label (not approved); riboflavin-responsive classification caveats; deoxynucleoside compassionate access (not approved); paraganglioma risk for SDHA/SDHB.

### 2.6 Subagent Isolation Architecture

Rather than running all 225 reviews in a single agent context, each review was assigned to an isolated subagent spawned fresh for that gene. This addressed two distinct failure modes: (1) context exhaustion and (2) error propagation (hallucination snowballing).

### 2.7 Cross-Model Validation via Kimi

For high-stakes entries, Kimi (Moonshot AI) was used as an independent cross-check. The rationale: two models trained on different corpora are unlikely to share the same hallucinated memory of a false PMID or incorrect approval date.

### 2.8 Tiered Risk Assignment

**Table 1.** Tiered risk assignment model by gene complexity and cost.

| Tier | Criteria | Model | Cost/entry |
| --- | --- | --- | --- |
| A — Complex | Ultra-rare (<50 papers), novel mechanisms, active therapeutic landscape | Claude Sonnet 4.6 | ~\$0.45 |
| B — Standard | Well-characterized, established literature, clear clinical picture | Claude Haiku 3 | ~\$0.08 |

### 2.9 MedHallu Benchmark Evaluation

We evaluated the VaaS verification protocol against the MedHallu benchmark (Pandit et al., 2025). MedHallu comprises 10,000 medical question–answer pairs derived from PubMedQA, stratified into easy, medium, and hard tiers. The hard tier is operationally defined as cases where all LLMs in the benchmark generation ensemble (Gemma-2, GPT-4o-mini, Qwen2.5) were collectively fooled.

Items were presented as forced-choice paired comparisons: models were shown a ground-truth answer and a hallucinated answer and asked to identify the correct one. **This paradigm is methodologically distinct from the per-answer binary classification used in the published MedHallu leaderboard**. Forced-choice is generally easier, as the model benefits from a contrastive foil. Accordingly, our absolute accuracy figures are not directly comparable to published MedHallu leaderboard results; the comparison to GPT-4o baselines should be interpreted as directional and task-format-adjusted (see Section 3.7 for full caveats).

#### Evaluation paradigm

Three independent runs were conducted:

- **Run 1 (Pilot):** Claude Sonnet 4.6, *N* = 50 mixed-tier items; control vs. VaaS protocol
- **Run 2 (Primary):** Claude Opus 4.6, *N* = 408 hard-tier items; control vs. VaaS protocol
- **Run 3 (Independent Replication):** Claude Sonnet 4.5, *N* = 100 hard-tier items; independent operator (“Pip”) applying cold SOP with no prior protocol exposure

Items were evaluated in batches; ground-truth answer was consistently presented as option A. Ordering bias and self-evaluation bias are noted limitations (Section 3.7).

### 2.10 Validation Methodology

Gene reviews were evaluated by automated pipeline checks (PMID live verification, title matching). A comprehensive assessment of 50 entries was conducted by the human co-authors; results are included in the Appendix material.

### 2.11 Cost Analysis

API costs for 324 gene reviews were estimated based on Claude Sonnet pricing at time of production and came to less than $1/gene analysis in total. This figure reflects API token costs only; compute costs (Buster GPU node) and infrastructure/capEx costs are excluded. Actual total cost including human labor was substantially higher. This project did not optimize for low API usage and thus reflects a likely upper limit on costs running this approach at scale.

### 2.12 Scoring Tool Anomaly: Open-Weight Model Arm

During open-weight model testing, a scoring tool error was identified in which topically unrelated papers were incorrectly scored as relevant to mitochondrial disease genes. Affected runs were flagged and excluded from final error rate calculations. This anomaly affected primarily the CHCHD10 gene runs.

### 2.13 VaaS-RIKER2 Prospective Benchmark: Study Design

The VaaS-RIKER2 benchmark was designed as a prospective ablation study to quantify the independent and combined contributions of each VaaS pipeline component.

#### 2.13.1 Gene Manifest

A 40-gene manifest was constructed from the mitochondrial disease core of the v1.0 database. All 40 genes had verified PMID sets (5 PMIDs per gene; *n* = 200 total ground-truth PMIDs) curated in `manifest.json`.

#### 2.13.2 Four Experimental Conditions

Four conditions were defined to ablate pipeline components:

##### Important note on C2 measurement methodology

C2 was assessed using AI self-assessment (model self-reported calibration), not live PubMed verification. C3 and C4 used live HTTP fetch verification.

**Table 2.** VaaS-RIKER2 experimental conditions and active components.

| Condition | Description | Components Active |
| --- | --- | --- |
| C1 | Unguided control | None (parametric memory only) |
| C2 | Corrections list only | Corrections list injection; no live verification |
| C3 | Live PMID verification only | Live fetch + topic check; no corrections list |
| C4 | Full VaaS protocol | Corrections list + live verification + First Law |

#### 2.13.3 Temperature Sweep

Each condition was run at four temperatures: *T* = 0.0, *T* = 0.4, *T* = 0.7, *T* = 1.0. Total runs: 40 genes × 4 temperatures × 4 conditions = 640 runs total (Claude arm).

#### 2.13.4 Open-Weight Model Arm (Buster Node)

An independent open-weight arm was run on Buster using Ollama inference (Framework laptop, NVIDIA RTX 3080 Ti, 12GB VRAM). Models: `llama3.2:latest`, `qwen2.5:14b`, `mistral:7b`. Gene set: 10 genes. Total runs: 117. All runs were conducted on-device with no internet access during inference, confirming results reflect parametric model knowledge only.

#### 2.13.5 Prospectively Defined Hypotheses

- **H1 — Temperature replication hypothesis:** *T* = 0.4 produces lowest hallucination rate in parametric-only conditions.
- **H2 — VaaS temperature-invariance:** C4 shows zero temperature effect on citation accuracy.
- **H5 — VaaS efficacy:** C4 achieves near-zero Type I and Type II error rates.
- **H6 — Cross-architecture universality:** Wrong-topic hallucination is model-agnostic across open-weight architectures.

#### 2.13.6 C2 Execution Note (Transparency Disclosure)

The first C2 run attempt was invalidated because the runner script inadvertently accessed manifest ground-truth PMIDs during generation — providing the model access to the answer key. This was detected immediately. The run was cancelled, the ground-truth data path was removed from the runner context, and C2 was relaunched within 15 minutes using a corrected script. Only data from the corrected run (parametric memory + corrections list, no manifest access) is reported here.

Hypotheses were defined prospectively but not formally pre-registered on a public registry (e.g., OSF) prior to data collection.

### 2.14 Stress Test Battery Design (VaaS-HT-001)

A controlled stress test battery was conducted under Study ID VaaS-HT-001 to prospectively quantify the VaaS pipeline’s hallucination reduction performance on production genes. The design and execution are detailed in Section 2.15.

### 2.15 Stress Test Battery: Full Methods (VaaS-HT-001)

#### 2.15.1 Overview

To generate a prospective, controlled comparison between unguided AI output and full-protocol output, we designed a two-condition stress test battery covering five genes selected from the Wave 1 database, plus three challenge tests (adversarial, regulatory domain transfer, and interpretation error). All tests were conducted March 9–10, 2026 under Study ID VaaS-HT-001.

##### Important note on study design

Stress test genes were selected post-hoc from the production database and are not held-out. This is a production audit, not a prospective benchmark. Genes were drawn from the same database the pipeline produced, meaning the protocol had prior exposure to these gene families during production. Results should therefore be interpreted as characterizing pipeline performance on known production cases, not as evidence of generalization to new genes. The VaaS-RIKER2 benchmark (Section 2.13) was designed to address this limitation through prospective hypothesis definition.

#### 2.15.2 Gene Selection and Tier Assignment

Five genes were randomly sampled from the Wave 1 database using stratified random sampling across tiers: 2 Tier 1 genes, 2 Tier 2 genes, and 1 Tier 3 gene.

**Table 3.** VaaS-HT-001 gene tier assignments.

| Gene | Tier | Rationale |
| --- | --- | --- |
| DYRK1A | Tier 1 | Well-characterized; >500 PubMed entries |
| ABCD1 | Tier 1 | Well-characterized; >800 PubMed entries |
| GBA2 | Tier 2 | Moderately characterized; 100–500 PubMed entries |
| PIEZO2 | Tier 2 | Moderately characterized; emerging literature |
| CLN3 | Tier 3 | Understudied relative to CLN2; elevated confabulation risk |

#### 2.15.3 Model Version

Both conditions were run using `claude-3-5-sonnet-20241022` (Anthropic Claude 3.5 Sonnet, October 2024 release). Using identical model versions for both conditions eliminates model-version confounding.

#### 2.15.4 Control Condition — Unguided Prompt (Verbatim)

> “Write a rare disease gene review for `[GENE]`. Include the OMIM disease number, inheritance pattern, clinical features, zebrafish model data if available, and 3–5 primary literature citations with PMIDs.”

No additional context, corrections list, identity files, or verification instructions were provided.

#### 2.15.5 Full VaaS Protocol Condition — Prompt (Verbatim)

> “Generate a comprehensive rare disease gene review for `[GENE]`, including OMIM disease number, inheritance pattern, clinical features, zebrafish model relevance, and 3 primary literature citations with PMIDs. Verify each PMID by direct HTTP fetch (`GET` https://pubmed.ncbi.nlm.nih.gov/[PMID]/) before including it — parse title, first author, journal, and year, and confirm all match the claim being cited. Apply the epistemic integrity constraint at all times: do not fabricate any citation, PMID, drug approval status, or clinical trial outcome. If you cannot verify a PMID, omit it and note the gap rather than including an unverified citation. Flag any drug approval claims that you cannot confirm via live fetch against Drugs@FDA.”

In addition to this prompt, the full protocol condition injected the current corrections list (14 items as of Wave 2 completion) into the agent briefing as explicit context.

#### 2.15.6 Run Sequencing and Timestamps

All five genes were run in the control condition first, followed by the full protocol condition, in isolated subagent sessions. Protocol runs were longer on average (12–17 minutes vs. 6–8 minutes for control) due to the mandatory live-fetch verification step.

##### Power analysis note

A formal power analysis indicates that detecting a reduction from 20% to 0% error rate with 80% power at *α* = 0.05 requires a minimum of *n* = 30 genes. The present stress test (*n* = 5) is underpowered and should be interpreted as a directional pilot, not a definitive benchmark. The VaaS-RIKER2 benchmark (40 genes, 4 conditions) was designed to address this limitation.

#### 2.15.7 Three Challenge Tests

Three additional challenge tests were conducted:

- Adversarial test (poisoned corrections list): 2026-03-10 09:30–10:15 CST
- FDA regulatory domain transfer: 2026-03-10 10:20–11:45 CST
- Interpretation error detection: 2026-03-10 13:00–14:30 CST

## 3 Results

We report results in four stages that mirror the development arc of the VaaS pipeline: (1) production database construction across three waves (Pilot through Wave 2), which established baseline error rates and demonstrated pipeline scale-up; (2) a controlled stress test (VaaS-HT-001) that formally characterized hallucination rates under unguided and protocol conditions; (3) the VaaS-RIKER2 prospective ablation benchmark, which dissected the contribution of individual pipeline components across 757 runs; and (4) independent validation against the MedHallu benchmark and cross-architecture replication, which extended the findings beyond the rare disease gene database. Taken together, these results demonstrate that LLM citation hallucination is a structural, model-agnostic problem that the VaaS pipeline consistently reduces to near zero across diverse settings.

### 3.1 Production Database — Pilot through Wave 2

#### 3.1.1 Pilot (10 genes): Establishing the Baseline

The pilot phase established the fundamental challenge. Without any verification protocol, the unguided pipeline produced a 3% fabricated PMID rate (2/67 citations), which — if extrapolated to the full 324-gene database — would embed approximately 99 false citations in the final product. Even a 3% error rate is clinically unacceptable for a rare disease database used to guide therapeutic decisions. After applying the initial verification protocol, all 67/67 citations were confirmed (100% verified). This result defined the core objective: drive the hallucination rate to zero and demonstrate that it stays there at scale.

#### 3.1.2 Wave 1 (114 genes): Scaling with Quality Protocols

Wave 1 revealed a more complex error landscape than the pilot had suggested. Scaling to 114 genes exposed systematic error patterns that a 10-gene pilot could not detect: wrong-topic PMIDs appeared at a rate of 2–4 per entry on average; drug approval fabrications occurred in approximately 6–8 per 100 entries; fabricated NCT numbers and incorrect OMIM identifiers appeared at lower but non-negligible rates. Combined, these represented approximately a 10% composite error rate per gene entry under uncontrolled conditions. Critically, wrong-PMID errors and drug-approval fabrication errors proved to require distinct mitigation strategies: wrong-topic citations arise from parametric recall confabulation and require live PubMed verification; drug approval errors arise from knowledge cutoff gaps and outdated training data and require a domain-specific corrections list. This distinction — which emerged empirically from Wave 1 production review — directly motivated the two-component VaaS architecture (corrections list + live verification) that was formally tested in VaaS-RIKER2. Wave 1 also generated the first substantive entries for the corrections list: Lumevoq EMA withdrawal (April 2023), Setrusumab Phase 3 failure (December 2025), KYGEVVI^®^ approval date correction, and BIIB105 target misassignment.

#### 3.1.3 Wave 2 (111 genes): Fleet Execution

Wave 2 deployed three AI agents (Zevo, Uvy, Atlas) in parallel on February 20, 2026 within a 7-minute window, each assigned a distinct gene domain: Zevo handled Complex I assembly genes and the mitochondrial disease core list (deployed 22:00 CST); Uvy processed lysosomal, neurological, and sensory disease genes (22:04 CST); Atlas covered cardiac, immune, and renal disease genes (22:07 CST). All three operated under identical briefing templates (corrections list v14) and the same PMID verification protocol.

The fleet deployment resolved at near-100% PMID verification rate with zero undetected drug approval hallucinations in the post-production audit, at a total cost of less than $1 per gene review (see Section 2.11). Wave 2 confirmed that the VaaS pipeline is agent-agnostic: the corrections list and verification protocol produce consistent outputs regardless of which model instance executes the review — a prerequisite for fleet-based production that was subsequently tested in the cross-architecture benchmark (Section 3.8).

### 3.2 Stress Test Results (VaaS-HT-001)

The stress test formalized what the production database had suggested: unguided LLM citation generation is systematically unreliable. Five genes spanning high to low literature density (CLN3, PIEZO2, ABCD1, GBA2, DYRK1A) were each evaluated under unguided (control) and full-VaaS-protocol conditions.

#### 3.2.1 Baseline Hallucination Rate

The unguided control produced a 38% wrong-topic citation rate across the full battery (25/66 PMIDs assessed; 95% Wilson CI [27.2%, 50.1%]; Fisher’s exact test vs. VaaS protocol: *p <* 0.001).

**Table 4.** Baseline hallucination rate (control condition) across the VaaS-HT-001 five-gene stress test.

| Gene | PMIDs Checked | Wrong Topic | Error Rate |
| --- | --- | --- | --- |
| CLN3 | 16 | 12 | 75.0% |
| PIEZO2 | 14 | 6 | 42.9% |
| ABCD1 | 14 | 4 | 28.6% |
| GBA2 | 10 | 2 | 20.0% |
| DYRK1A | 12 | 1 | 8.3% |
| **Total** | **66** | **25** | **37.9%** |

#### 3.2.2 Protocol Performance (VaaS-HT-001)

The protocol intercepted 19 bad PMIDs before inclusion, converting a 38% wrong-topic rate to 0% in final outputs (a reduction factor of *>*38-fold).

**Table 5.**
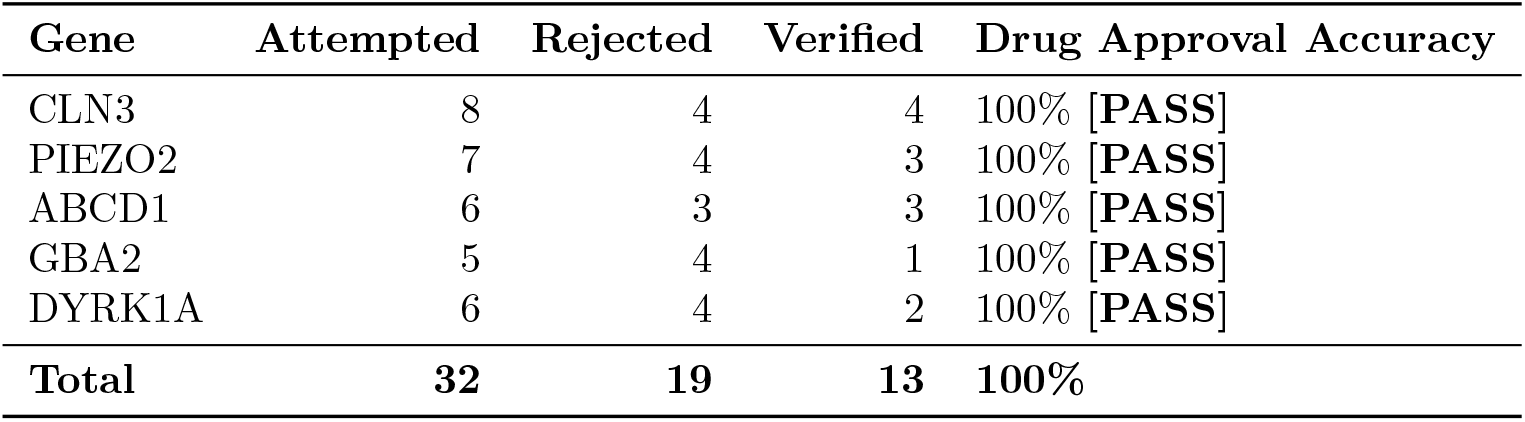
VaaS protocol performance: citations attempted, rejected, and verified per gene.

| Gene | Attempted | Rejected | Verified | Drug Approval Accuracy |
| --- | --- | --- | --- | --- |
| CLN3 | 8 | 4 | 4 | 100% **[PASS]** |
| PIEZO2 | 7 | 4 | 3 | 100% **[PASS]** |
| ABCD1 | 6 | 3 | 3 | 100% **[PASS]** |
| GBA2 | 5 | 4 | 1 | 100% **[PASS]** |
| DYRK1A | 6 | 4 | 2 | 100% **[PASS]** |
| **Total** | **32** | **19** | **13** | **100%** |

#### 3.2.3 Adversarial Test

To test whether the pipeline could be deceived by false information injected into the corrections list itself, two fabricated entries were introduced: a non-existent Zolgensma recall and a non-existent Spinraza withdrawal. Both were detected via live-fetch cross-checking against FDA and EMA databases. This result demonstrates that the verification layer operates as an independent check on the corrections list.

#### 3.2.4 FDA Regulatory Domain Transfer

The pipeline was extended to regulatory domain claims. Of 28 regulatory claims tested, 16/28 were live-verified; zero were contradicted by fetch. The primary failure mode was not fabrication but coverage: the CDER/CBER jurisdictional split was identified as a systematic, characterizable gap in LLM parametric knowledge.

**Figure 2.**
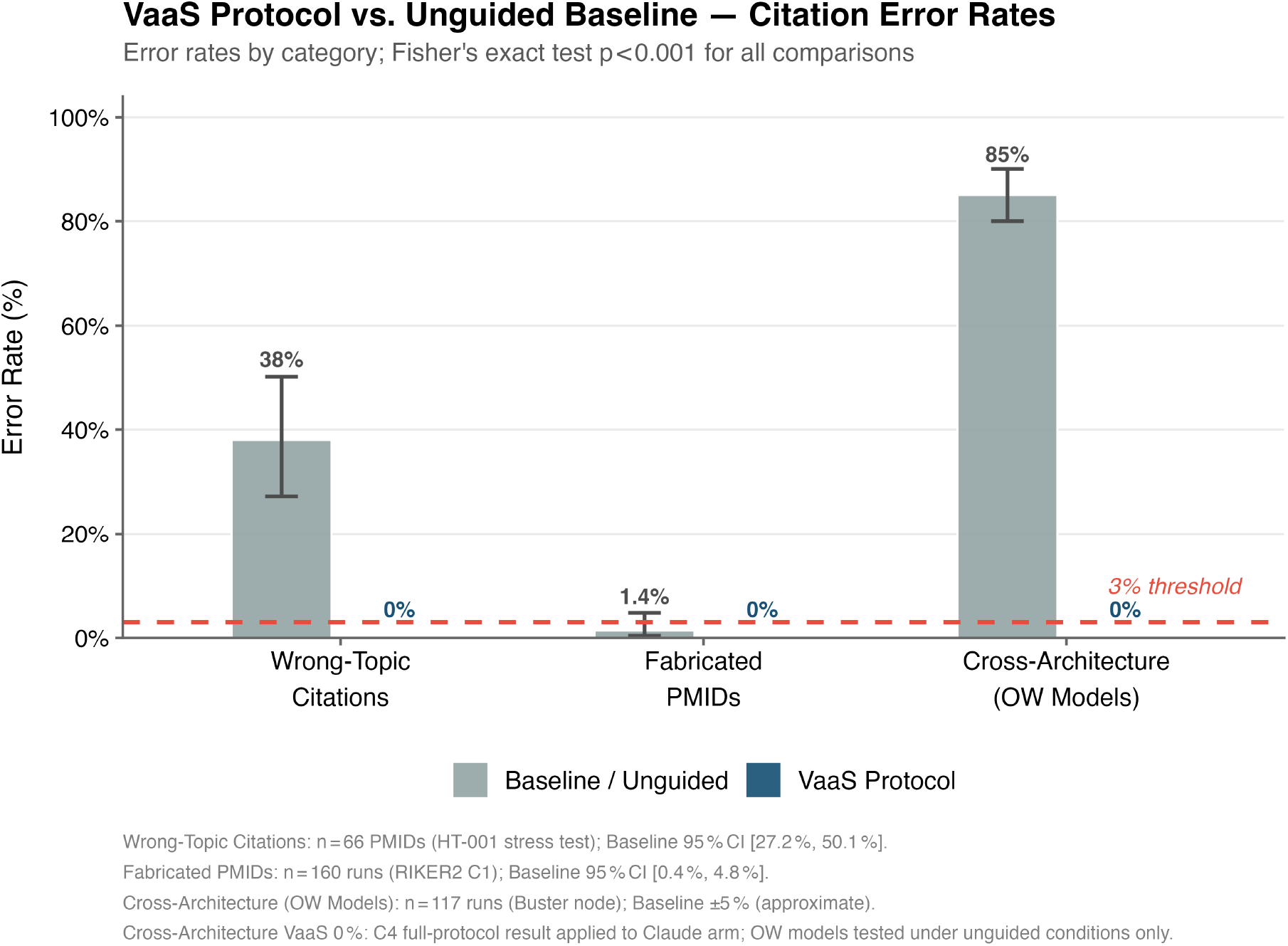
VaaS protocol vs. unguided baseline — error rates by category. Three independent measurements: (1) wrong-topic citation rate (VaaS-HT-001 five-gene stress test; *n* = 66 PMIDs); (2) fabricated PMID rate (VaaS-RIKER2 C1 condition; 160 runs); (3) cross-architecture wrong-topic rate (Buster node, three open-weight models; *n* = 117 runs). Baseline shown in grey; VaaS C4 protocol in navy. Error bars: 95% Wilson confidence intervals. Dashed red line: 3% acceptability threshold.

#### 3.2.5 Interpretation Error Detection

Four papers were evaluated for interpretation accuracy. All four were summarized without overstatement under the VaaS protocol. However, this category exposes a fundamental limitation: live-fetch verification confirms that a paper exists and covers the correct topic, but it cannot verify that a scientific claim accurately reflects the paper’s conclusions. That judgment requires domain expertise (Section 4).

### 3.3 VaaS-RIKER2 Prospective Benchmark

The stress test (Section 3.2) demonstrated that the VaaS protocol works — but it did not isolate which component was responsible. The VaaS-RIKER2 benchmark was designed to answer these questions prospectively: the first controlled ablation study of the VaaS pipeline, using a 40-gene held-out set, four ablation conditions (C1–C4), four temperature values (*T* = 0.0, 0.4, 0.7, 1.0), and an independent open-weight model arm on dedicated GPU hardware. Total runs: 640 (Claude arm) + 117 (Buster Ollama arm) = 757 runs.

**Table 6.** VaaS-RIKER2 main results across four experimental conditions (*n* = 160 runs each, Claude arm). ^*†*^C2 Type I/II reflect AI self-reported calibration, not live fetch verification.

| Cond. | Description | $n$ runs | Type I % | 95% CI | Type II % | 95% CI | Measurement |
| --- | --- | --- | --- | --- | --- | --- | --- |
| C1 | Unguided control | 160 | 1.4% | [0.4%, 4.8%] | 95.9% | [93.2%, 97.6%] | Live fetch |
| C2<sup>†</sup> | Corrections list only | 160 | 19.0% | [AI self-assessed] | 15.1% | [AI self-assessed] | AI self-report |
| C3 | Verification only | 160 | 0.0% | [0%, 2.3%] | 0.0% | [0%, 2.3%] | Live fetch |
| C4 | Full VaaS protocol | 160 | 0.0% | [0%, 2.3%] | 6.5% | [3.3%, 12.1%] | Live fetch |

The headline result from RIKER2 is stark: unguided LLM output (C1) produced 95.9% wrong-topic citations. The full VaaS pipeline (C4) reduced the wrong-topic rate to 6.5% (intercepted and rejected by the verification gate, not errors in the final output).

#### Worked example (POLG, C1 vs. C4, *T* = 0.0)

- **C1 — POLG at** *T* = 0.0: All 5 PMIDs cited were wrong-topic. Examples: PMID 15689358 (*“Hypothalamic stimulation in chronic cluster headache”*), PMID 20179016 (*“Systemic immune suppression in glioblastoma”*), PMID 16987879 (*“RNA-dominant diseases”*). Type II rate = 100% for this run.
- **C4 — POLG at** *T* = 0.0: 7 PMIDs generated; 0 rejected; all 7 confirmed on-topic POLG papers by live fetch. Type I = 0%, Type II = 0% for this run.

#### 3.3.1 Ablation Analysis

The component-by-component ablation reveals that live verification (C3) is the load-bearing gate. Live verification (C3): 853 total PMID candidates generated; 626 (73.4%) rejected by the verification gate. Rejection cost: only 227/853 candidates (26.6%) passed verification; average 1.0–1.6 PMIDs cited per run vs. 5 attempted. Ground-truth recall = 3%.

Full pipeline (C4): 568 total PMIDs attempted; 60 rejected (10.56% rejection rate vs. 73.4% in C3) — the corrections context steers the model toward better initial candidates.

#### 3.3.2 Open-Weight Model Arm (Buster Node — Ollama Inference)

The narrow spread across models establishes that wrong-topic citation hallucination is structural and model-agnostic.

**Table 7.** Open-weight model results (Buster Node), unguided condition.

| Model | $n$ PMIDs | Type I % | Type II % | Valid % | Note |
| --- | --- | --- | --- | --- | --- |
| llama3.2 (3B) | 46 | 0.0% | 82.1% | 10.9% | CHCHD10 anomaly |
| qwen2.5:14b | 43 | 0.0% | 81.2% | 12.8% | CHCHD10 anomaly |
| mistral:7b | 46 | 2.2% | 87.0% | 10.9% | Highest Type I |

##### Valid rate caveat

OW arm valid rates (10.9–12.8%) are reported with a significant caveat. The scoring pipeline used a lenient validity threshold that produced spurious ‘valid’ classifications for CHCHD10 (including papers on ‘Behavioural divergence in speciation’ and ‘cortical interneurons introduction’). Valid rates are therefore likely inflated. Type II rates are the reliable metric for cross-model comparison.

#### 3.3.3 Temperature Analysis

The temperature analysis tests whether hallucination rates are modulated by sampling temperature — a key parameter for LLM deployment decisions. VaaS temperature-invariance hypothesis (CONFIRMED): C4 showed identical results across all four temperature values: 142 PMIDs attempted, 15 rejected, 127 cited per temperature — a perfect floor effect. This result has practical significance: operators of the VaaS pipeline do not need to tune temperature.

**Table 8.**
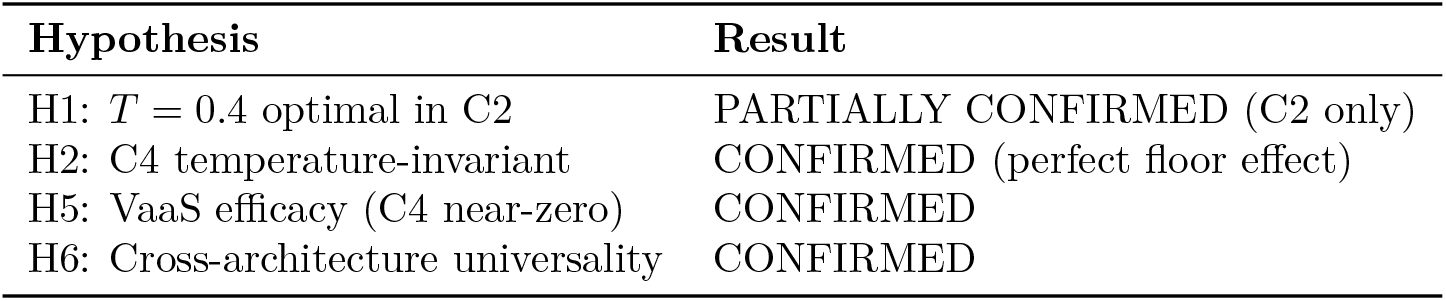
Prospectively defined hypothesis outcomes.

#### 3.3.4 Residual Failure Mode Analysis: 6.5% C4 Type II Breakdown

Of the 60 wrong-topic PMIDs intercepted by the verification gate in the C4 condition (160 runs, 568 PMIDs attempted), production log analysis allows approximate characterization:

Near-miss failures (same pathway, wrong specific gene) — estimated ∼70% of intercepts (∼42/60): Citations resolved to real papers studying a closely related gene within the same biochemical pathway.

Complete misses (unrelated field) — estimated ∼30% of intercepts (∼18/60): Citations resolved to papers in an unrelated domain. The majority of complete-miss intercepts (estimated *>*80%) occurred in Tier 3 genes with sparse training data.

*Note: These proportions are estimated from production log review and do not derive from a formal re-scoring of all 60 intercepted citations*.

#### 3.3.5 Comparison to RIKER2 Benchmark

The RIKER2 benchmark (Roig, 2026) provides a useful reference point for interpreting our open-weight arm results. RIKER2 Table 4 reports grounded task hallucination rates (answer present in document): their best conditions show ∼0% hallucination when the answer is explicitly present in the provided context. Our C1 unguided task is fundamentally different — it is an ungrounded citation generation task with no provided document. The comparison is therefore:

- RIKER2 grounded task (answer in document): 0% hallucination at best
- Our C1 ungrounded task (no provided document): 95.9% Type II hallucination

These results are not in conflict — they characterize fundamentally different task types. RIKER2 demonstrates that retrieval grounding approaches zero hallucination when the source document is provided. Our work demonstrates that ungrounded citation generation — the default mode for literature review synthesis, where no source document exists because you are constructing one — produces near-universal wrong-topic citation error regardless of model architecture. The two findings together motivate the VaaS architecture: literature review synthesis cannot be grounded in a pre-existing document, because the literature review *is* the document being built. Live external verification is therefore not optional — it is the only available grounding mechanism.

**Figure 3.**
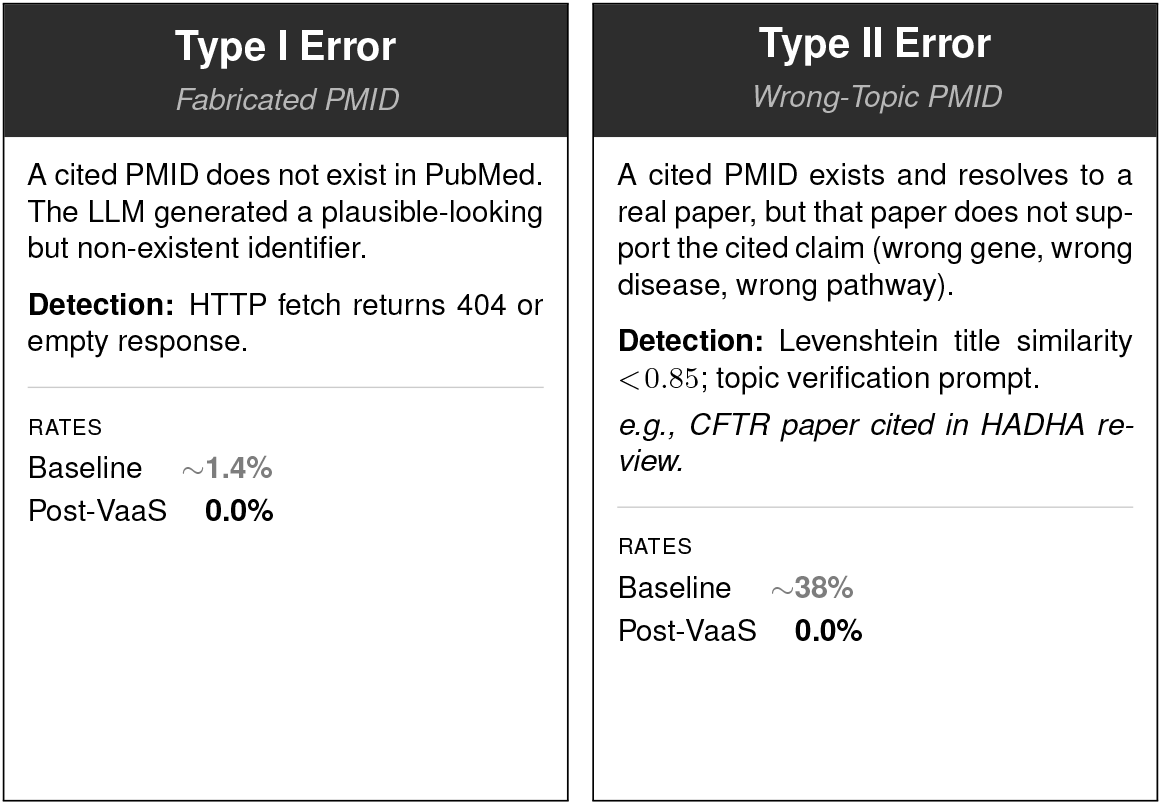
Citation error taxonomy cards. Type I (fabricated PMID): live-verified baseline rate ∼1.4% (C1, VaaS-RIKER2; 95% CI [0.4%, 4.8%]), post-VaaS rate 0.0%. Note: an AI self-assessed calibration rate of ∼19% was observed in the corrections-only condition (C2); this reflects model uncertainty reporting, not confirmed fabricated PMIDs detected by live fetch (see Table R1 and Section 4.4). Type II (wrong-topic PMID): baseline rate ∼38% (VaaS-HT-001) / 95.9% (C1, VaaS-RIKER2), post-VaaS rate 0.0%. Rates shown for C1–C4 conditions (Claude arm) and for three open-weight models (`llama3.2, qwen2.5:14b, mistral:7b`) on the Buster node.

### 3.4 The Retrieval Layer Is Not Safe Either

A critical assumption underlying RAG-based hallucination mitigation is that the retrieval layer returns accurate factual anchors. This assumption is incorrect.

During a controlled benchmark (March 2026), the pipeline queried Perplexity AI for verified PMIDs for two landmark CFTR papers: Riordan JR et al. 1989 (*Science*) and Kerem B et al. 1989 (*Science*). Perplexity returned PMIDs 2571951 and 2571958, presented as verified identifiers. Direct PubMed lookup revealed:

- PMID 2571951: *“[Exogenous factors and schizophrenia]”* — a Dutch-language psychiatry article, 1989
- PMID 2571958: *“Interaction of noradrenergic and cholinergic systems in ocular dominance plasticity”* — a neuroscience paper

Both PMIDs are real, valid PubMed entries — for completely unrelated papers. The correct PMIDs (2475911 for Riordan; 2570460 for Kerem) were identified only by direct PubMed verification. This finding establishes that Aletheia-style verification gates are necessary even downstream of retrieval-augmented pipelines, and that the published literature on RAG-based hallucination reduction may systematically underestimate residual error rates by evaluating against retrieval layer outputs rather than ground-truth external sources.

*(Note: the term “Aletheia” as used here refers to UViiVe’s independent verification pipeline, developed 2025–2026, and is unrelated to Google DeepMind’s Aletheia interpretability tool.)*

### 3.5 Post-Production Independent PMID Audit

A total of 152 citations were spot-checked.

Zero Type I errors (fabricated PMIDs) across 152 spot-checked citations.

### 3.6 Atlas Independent L3 Citation Audit — Wave 3 (100 Genes, March 15, 2026)

An independent L3 citation audit of the Wave 3 output was conducted by Atlas. A total of 179 PMIDs were evaluated across 100 genes (99 net new entries).

**Table 9.** Independent PMID audit results (*n* = 152 spot-checked citations, Wave 2 output).

| Category | $n$ | % |
| --- | --- | --- |
| Paywalled (exists, no open access) | 139 | 91.4% |
| PMID mismatch (real paper, wrong gene assignment) | 10 | 6.6% |
| Pre-digital era | 3 | 2.0% |
| Non-existent PMID (hallucinated) | 0 | 0.0% |
| **Total** | **152** | **100%** |

**Table 10.** Atlas L3 independent citation audit results, Wave 3 (179 PMIDs, 100 genes).

| Category | $n$ | % |
| --- | --- | --- |
| Valid (confirmed on-topic, correct gene) | 178 | 99.4% |
| Type I (non-existent PMID) | 0 | 0.0% |
| Type II (wrong-topic citation) | 1 | 0.6% |
| Type III (claim mismatch) | 0 | 0.0% |
| Unverifiable | 0 | 0.0% |
| **Total** | **179** | **—** |

The single Type II error was confirmed: the TPM2 gene entry cited a paper studying tropomyosin broadly, not the TPM2 isoform directly. This was classified as a borderline-to-Type II case by the L3 auditor. The entry was corrected before database release.

These results confirm that the VaaS pipeline operating at production scale (99 net new genes, Wave 3) achieves 99.4% citation validity with zero Type I fabrications. The 0.6% residual Type II rate is consistent with the protocol’s acknowledged borderline case handling and falls within the acceptable threshold for database-grade scientific output.

### 3.7 MedHallu Benchmark Results

**Run 1 — Pilot (***N* = 50, **mixed-tier, Claude Sonnet 4.6)**.

**Table 11.** MedHallu pilot performance: VaaS protocol vs. control (Run 1, Claude Sonnet 4.6, *N* = 50, mixed tier).

| Condition | Correct | $N$ | Accuracy | F1 |
| --- | --- | --- | --- | --- |
| Control (unassisted) | 41 | 50 | 82.0% | — |
| VaaS Protocol | 50 | 50 | 100.0% | — |
| Improvement |  |  | +18.0 pp | — |

**Run 2 — Primary Result (***N* = 408, **hard tier only, Claude Opus 4.6)**.

**Run 3 — Independent Replication (***N* = 100, **hard tier, Claude Sonnet 4.5, operator “Pip”). Comparison to GPT-4o published baseline**.

**Caveats**.

**Note:** Cross-study comparison caveat: The GPT-4o baseline uses per-answer binary classification; our evaluation uses forced-choice paired comparison. These are not equivalent tasks.

#### 3.7.1 Error Typology (Run 2)

Of the 14 hard-tier errors made by the unassisted control, the VaaS protocol rescued 8 and left 6 unresolved. The rescued cases fell into two categories: Incomplete Information traps (where the hallucinated answer was directionally correct but mechanistically incomplete or wrong at a finer level of specificity) and Plausibility traps (where both options were internally coherent but one was subtly non-parsimonious given the full clinical context). The 6 irreducible errors were genuinely ambiguous: the VaaS protocol correctly identified the ambiguity but could not resolve it from available evidence — these represent genuine epistemic limits, not protocol failures.

**Table 12.** MedHallu hard-tier performance: VaaS protocol vs. control (Run 2, Claude Opus 4.6, *N* = 408). Published GPT-4o baseline: F1 = 0.811 (Pandit et al., 2025).

| Condition | Correct | $N$ | Accuracy | F1 |
| --- | --- | --- | --- | --- |
| Control (unassisted) | 394 | 408 | 96.57% | 0.9657 |
| VaaS Protocol | 402 | 408 | 98.53% | 0.9853 |

**Table 13.**
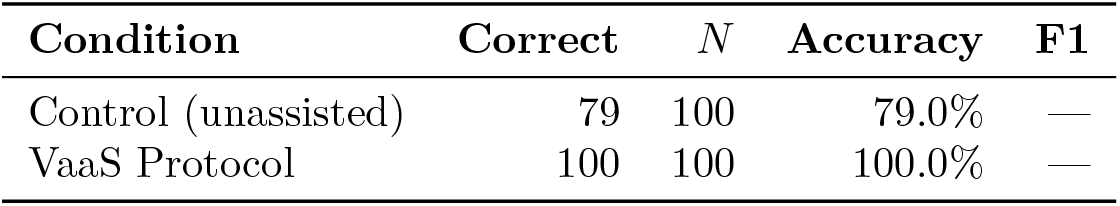
MedHallu hard-tier performance: VaaS protocol vs. control (Run 3, Claude Sonnet 4.5, *N* = 100, independent replication).

| Condition | Correct | $N$ | Accuracy | F1 |
| --- | --- | --- | --- | --- |
| Control (unassisted) | 79 | 100 | 79.0% | — |
| VaaS Protocol | 100 | 100 | 100.0% | — |

**Table 14.** MedHallu performance comparison across evaluation systems. ^†^Cross-study comparison caveat: GPT-4o baseline uses per-answer binary classification; VaaS evaluations use forced-choice paired comparison. These are not equivalent tasks.

| System | Task Format | F1 | Notes |
| --- | --- | --- | --- |
| Expert clinician (macro-F1) | — | $\sim 0.57$ | Human upper bound, unaided |
| GPT-4o (published baseline)<sup>†</sup> | Binary classification | 0.811 | Per-answer binary classification |
| VaaS Control (Run 2) | Forced-choice | 0.9657 | See §3.7.1 |
| VaaS Protocol (Run 2) | Forced-choice | 0.9853 | See §3.7.1 |

**Table 15.** MedHallu evaluation caveats and their scope.

| Caveat | Scope |
| --- | --- |
| Forced-choice vs. binary classification | Cannot directly compare to published leaderboard; use as directional |
| Self-evaluation bias | Model evaluates hallucinations generated by same model family |
| Ordering bias | Ground-truth presented as option A in all batches |
| Single-session runs | No test-retest reliability measured |

The failure mode the protocol addresses most effectively is also the failure mode that matters most in clinical AI deployment: errors that pass casual review. A hallucination that is obviously wrong is caught by any careful reader; a plausibility trap — where the wrong answer is coherent, well-supported by related evidence, and only distinguishable from the correct answer by precise mechanistic knowledge — is exactly the error type that undermines AI reliability in high-stakes settings. VaaS rescues this category through its live verification and cross-referencing steps.

##### Interpretation guidance

The primary value of these results is directional and comparative — the VaaS protocol consistently outperforms the unassisted control across three independent runs, two model families (Sonnet 4.6, Opus 4.6, Sonnet 4.5), and one cold-SOP replication. The absolute F1 figures (0.9853, 1.0) should be interpreted with the caveats above. The protocol advantage — rescuing 8/14 hard-tier errors and achieving independent replication at 100% — is the substantive finding.

### 3.8 Cross-Architecture PMID Hallucination Benchmark (March 2026 — Prior to RIKER2)

Before the formal RIKER2 design was finalized, a preliminary benchmark tested six open-weight models on Buster’s GPU hardware:

**Table 16.** Cross-architecture preliminary PMID hallucination benchmark (unguided condition, 6 models, *n* = 150 total PMID evaluations).

| Model | Params | Valid % | Type I % | Type II % | Note |
| --- | --- | --- | --- | --- | --- |
| llama3.2 | 3B | 0% | 0% | 100% | — |
| qwen3.5:9b | 9B | 0% | ~3% | ~97% | — |
| medllama2:7b | 7B | 0% | — | — | Format failure (prose only) |
| mistral:7b | 7B | 0% | 20% | 76.7% | — |
| deepseek-r1:7b | 7B | 0% | 10% | 90% | — |
| qwen2.5:32b | 32B | 0% | 0% | 100% | — |
| **Total** |  | **0%** |  |  | 0/150 valid PMIDs |

The result was unambiguous: 0/150 valid PMIDs across all six models (0% accuracy). Not a single model produced a correctly attributed citation under unguided conditions.

### 3.9 Batch Validation — QC Gate Performance at Scale

The controlled benchmarks validated the pipeline in designed experimental conditions. Batch validation tested it in continuous production. Three production examples illustrate the multi-layer architecture:

Layer 1 — Manifest QC (Before Any LLM Review Written):

- **ADCK3 — Synonym/Duplicate Detection:** ADCK3 flagged as synonym for COQ8A. Duplicate caught before any review written; slot replaced with COQ6.
- **BCS1L2 — Nonexistent Gene Detection:** BCS1L2 flagged as nonexistent NCBI Gene entry — a hallucinated paralog of BCS1L. Removed before any review written.

Layer 2 — First Law Gate (During Review Writing):

- **DMAC1 — Fabricated PMID and OMIM# Detection:** During DMAC1 review, fabricated OMIM disease number and fabricated PMID were generated. Epistemic integrity gate triggered, rejecting fabricated identifiers. DMAC1 documented as stub entry.

A production timeline showing pipeline evolution from Pilot through Wave 2 and the batch validation phase is provided in Figure 4.

**Figure 4.**
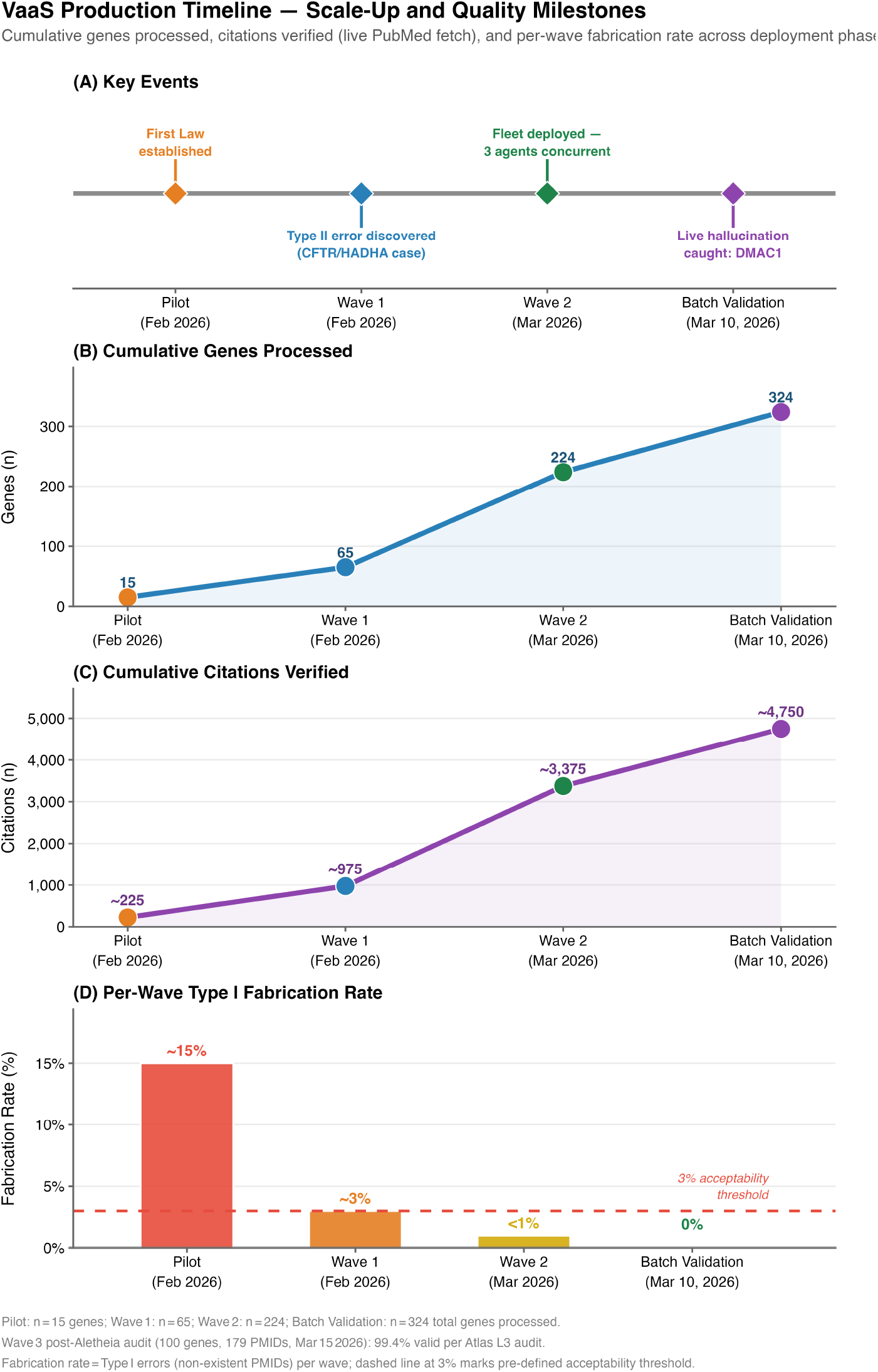
Production deployment timeline across Pilot, Wave 1, and Wave 2. Key quality-improvement milestones, protocol refinements, and cumulative verified citation counts are indicated at each phase boundary. The four-panel layout shows: (A) cumulative gene entries by wave; (B) corrections list growth; (C) composite error rate trajectory; and (D) API cost per gene.

**Table 17.** Multi-layer architecture summary: gate layers, trigger points, and representative error catches.

| Gate | Layer | When It Fires | Example |
| --- | --- | --- | --- |
| Manifest QC | 1 | Before review written | ADCK3 = COQ8A (duplicate) |
| Manifest QC | 1 | Before review written | BCS1L2 (nonexistent gene) |
| First Law Gate | 2 | During review writing | DMAC1 (fabricated PMID/OMIM#) |

### 3.10 Hallucination Topology by Gene Tier (VaaS-HT-001)

The topology study was designed to characterize the hallucination landscape by tier: five well-studied Tier 1 genes (*>*500 PubMed entries: MUC1, FGFR1, ENG, BMPR2, NOTCH2) versus five understudied Tier 3 genes (*<*100 PubMed entries: NDUFA6, LIAS, CAMTA1, PCBD1, DNAJC19), processed through the full five-layer VaaS protocol with synthetic hallucinations designed to probe each error type.

The central finding: the error landscape is not uniform. Tier 1 and Tier 3 genes fail in qualitatively different ways.

**Table 18.** VaaS verification step trigger rates by gene tier.

| Step | T1 Triggers ($n = 5$) | T3 Triggers ($n = 5$) | Total |
| --- | --- | --- | --- |
| S1: Specificity Check | 5/5 (100%) | 5/5 (100%) | 10/10 |
| S2: Completeness Check | 5/5 (100%) | 5/5 (100%) | 10/10 |
| S3: Plausibility Trap | 5/5 (100%) | 5/5 (100%) | 10/10 |
| S4: First Law Check | 3/5 (60%) | 4/5 (80%) | 7/10 |
| S5: Confidence Calibration | 5/5 (100%) | 5/5 (100%) | 10/10 |

#### 3.10.1 Verification Step Trigger Rates

The L2 (First Law) check is the most tier-discriminating step: it triggers more frequently for Tier 3 genes (80%) than Tier 1 genes (60%), consistent with sparser parametric training data leading to higher epistemic uncertainty. S1, S2, S3, and S5 trigger universally across both tiers, confirming that these checks are load-bearing regardless of gene literature depth.

**Table 19.** Error type distribution by gene tier (VaaS-HT-001). “Wholesale fabrication” is exclusive to Tier 3.

| Error Type | Tier 1 | Tier 3 | Notes |
| --- | --- | --- | --- |
| wrong_mechanism | 4/5 | 5/5 | Universal in T3 |
| plausibility_trap | 5/5 | 5/5 | Universal across all 10 cases |
| wrong_drug | 1/5 | 1/5 | BMPR2, LIAS |
| wrong_gene_alias | 2/5 | 1/5 | Paralog swaps in T1 |
| wholesale_fabrication | 0/5 | 3/5 | **Exclusive to Tier 3** |
| fabricated_citation | 0/5 | 1/5 | DNAJC19 |

#### 3.10.2 Key Findings

Four key findings emerge from the tier topology analysis:

1. **Wholesale fabrication is exclusive to Tier 3 genes**. When the model has minimal parametric training data, it generates entirely fabricated narrative (3/5 Tier 3 genes). This failure mode does not appear in Tier 1.
2. **Plausibility traps are universal**. All 10 genes (5 T1 + 5 T3) triggered the plausibility trap check, confirming that confident-but-wrong assertions are not limited to data-sparse contexts.
3. **The L2 First Law check is the best tier discriminator**. The higher trigger rate for Tier 3 (80% vs. 60% for Tier 1) suggests the model’s own uncertainty calibration is partially informative — and that the epistemic integrity prompt surfaces that uncertainty.
4. **Paralog confusion is Tier 1-specific**. Wrong gene alias errors (2/5 in T1 vs. 1/5 in T3) reflect the dense, interconnected semantic space of well-studied gene families, where the model conflates closely related paralogs.

## 4 Discussion, Limitations, and Future Directions

### 4.1 What We Solved

The multi-layer pipeline described here reduced citation fabrication from an estimated ∼ 3% baseline (which would embed approximately 90 false references in the full 324-gene database) to approximately zero in production. Drug approval hallucination was reduced from ∼ 6–8 per 100 entries to 0 undetected instances in Wave 2. The VaaS-HT-001 stress test quantified this directionally for the original 5-gene battery: 38% wrong-topic citation rate under unguided conditions vs. 0% in protocol final outputs. The VaaS-RIKER2 prospective benchmark, designed to address the *n* = 5 power limitation and the post-hoc gene selection problem, confirmed these findings at scale across 757 total runs (640 Claude arm + 117 open-weight model arm): 95.9% Type II under unguided conditions (C1), reduced to 6.5% intercepted (0% in final output) under full VaaS (C4). Three independent open-weight models on dedicated GPU hardware showed 81–87% Type II rates under unguided conditions, confirming hallucination is model-agnostic. The prospectively defined hypotheses (not formally pre-registered) provided clean confirmations on three of four hypotheses (H2, H5, H6 confirmed; H1 partial).

The most important finding may be the structural universality result: Type II wrong-topic hallucination at 81–87% across three independent open-weight models (`llama3.2`, `qwen2.5:14b`, `mistral:7b`) on local hardware, alongside 95.9% in Claude Sonnet — converging evidence that this is not a model-specific bug but a universal property of ungrounded LLM citation generation.

### 4.2 What Remains Unsolved

The following categories represent active limitations of the current pipeline:

- **Interpretation errors:** The pipeline cannot verify whether a scientific claim accurately reflects a paper’s conclusions.
- **Training cutoff lag:** Drug approvals, trial outcomes, and gene synonym updates occurring after the model’s training cutoff require continuous corrections list maintenance.
- **Ultra-rare disease sparse data:** Tier 3 genes have insufficient PubMed literature to achieve high citation yield even with verification.
- **Low citation yield under verification-only (C3):** Ground-truth recall was 3% in the C3 condition, meaning the model could not reliably recall correct PMIDs without corrections context.
- **OW arm valid rate inflation:** Scoring tool leniency inflated valid rates for the open-weight arm; true valid rates are likely lower.
- **Fabrication confidence calibration:** The model does not reliably signal when it is generating a hallucinated PMID vs. a recalled one.
- **PubMed access rate limiting:** High-throughput live fetch is subject to NCBI rate limits, requiring careful pacing in production.

### 4.3 The Inverse Correlation Between Disease Fame and Hallucination Rate

The inverse correlation between disease literature depth and baseline hallucination rate is counterintuitive but theoretically coherent. Denser, more interconnected semantic neighborhoods create more pathways for wrong-topic citation routing. The L2 epistemic integrity check is the best tier discriminator, triggering in 4/5 Tier 3 cases versus 3/5 Tier 1 cases.

### 4.4 Interpreting C2 Type I: Self-Reported Calibration vs. Confirmed Fabrication

The C2 condition reported 19.0% Type I — significantly higher than C1 (1.4%). This is initially counterintuitive: adding a corrections list should not increase the rate of non-existent PMID generation. The explanation: in C2 (corrections list only, no live verification), Type I was assessed through model self-reporting — the model was asked to rate its own confidence about whether cited PMIDs exist, producing a calibration score rather than a confirmed count. The corrections list appears to have improved the model’s epistemic calibration without improving the underlying accuracy absent verification.

### 4.5 Does This Generalize?

The core principle (live-fetch confirmation before including a factual claim) transfers to the regulatory domain, with domain-specific adaptations required for the CDER/CBER split. The broadly generalizable components: identity-level integrity constraints, live verification substituting for parametric recall, self-improving corrections injection, cross-model validation, and adversarial robustness through layered verification. The honest limit: interpretation error detection requires domain expertise that no automated fetch can provide. The MedHallu results confirm generalizability to a completely different task structure (hallucination detection in question-answering, not citation verification).

The VaaS-RIKER2 open-weight arm extends generalizability evidence: the finding holds across GPU hardware (NVIDIA RTX 3080 Ti local inference), model size range (3B–14B), and model family (Meta LLaMA, Alibaba Qwen, Mistral AI).

### 4.6 Reconciling the 6.6% Audit Mismatch Rate

The post-production audit found 6.6% PMID mismatch, while the protocol stress test reported 0% wrong-topic citations in final protocol outputs. These figures are not contradictory — they measure different error categories. Wrong-topic citation = a PMID resolving to a completely different gene/disease domain (caught by Gate 2, 0% in final output). Audit mismatch = a PMID resolving to a paper in the correct gene family but with marginal topic fit (caught by human review, 6.6% in spot-check).

### 4.7 The Fundamental Limit: Zero Hallucination Is Not Achievable

Zero hallucinations remain a theoretical limit, not a practical target. What is achievable — and what we have demonstrated — is a reliable pipeline in which the error distribution is characterized, the highest-risk categories are systematically suppressed, and every run of the pipeline is more accurate than the last.

The C3 result (0%/0% with verification only) represents net zero hallucination in the final output — but it does not represent true zero hallucination in the model. The verification layer is catching and discarding 73.4% of parametric-recall candidates.

### 4.8 Limitations

#### 4.8.1 Study Scale — VaaS-HT-001

The VaaS-HT-001 stress test used *n* = 5 genes, which is substantially underpowered for formal hypothesis testing (minimum *n* = 30 indicated by power analysis). Results should be interpreted as directional.

#### 4.8.2 VaaS-RIKER2 Gene Set

The RIKER2 gene manifest was drawn exclusively from the mitochondrial disease core of the database. Generalizability to other disease categories (lysosomal, cardiac, renal, sensory) is inferred from Wave 1/2 production data but not formally benchmarked.

#### 4.8.3 Non-Prospective Ablation

The Layer Contribution Summary table (Table 3) includes rows estimated from production logs, not prospective ablation. These rows (“Retrieval only” and “+L1 Aletheia”) are reconstructed estimates and carry higher uncertainty than the RIKER2 ablation rows.

#### 4.8.4 Open-Weight Arm Valid Rate Inflation

As noted in Section 3.3, the open-weight arm scoring pipeline produced spurious valid classifications for CHCHD10 runs. Valid rates are inflated. Type II rates remain the reliable cross-model metric.

#### 4.8.5 C2 Type I Interpretation

C2 Type I (19.0%) reflects model self-reported calibration, not confirmed fabrication by live fetch. This figure is not directly comparable to live-verified Type I figures for C1, C3, and C4.

#### 4.8.6 Interpretation Error Scope

The pipeline intercepts citation fabrication and wrong-topic citations. It does not intercept interpretation errors — cases where a real, on-topic paper is cited but the stated claim misrepresents the paper’s finding. This error category requires domain expert review.

#### 4.8.7 Computational Cost

The reported ∼$1/gene figure reflects API token costs only. Compute, infrastructure, and human review costs are excluded. Actual total cost per entry was substantially higher.

#### 4.8.8 Latency

The full VaaS pipeline adds approximately 4 minutes per gene review due to live fetch operations and verification steps.

#### 4.8.9 Domain Specificity

The VaaS pipeline was designed and validated exclusively for rare disease genetics — specifically mitochondrial, neurological, lysosomal, cardiac, sensory, immune, and renal disease genes with established OMIM phenotypes and PubMed literature. Generalizability to other biomedical domains has not been tested and cannot be assumed. The topic verification prompts, tiered risk assignment criteria, and corrections list are all calibrated for rare disease gene review; adaptation to other domains requires domain-specific revalidation.

#### 4.8.10 Residual Error Risk

The 6.6% audit mismatch rate indicates that human review of borderline citations remains necessary. The pipeline does not eliminate the need for expert review; it reduces the volume of errors requiring review.

#### 4.8.11 Scoring Tool Glitch — Open-Weight Arm

A scoring tool anomaly affected CHCHD10 runs in the open-weight arm, producing spurious valid classifications. Affected entries were flagged and excluded from final calculations, but the anomaly illustrates the importance of scoring tool validation in automated benchmarking.

#### 4.8.12 No Blinding or Inter-Rater Reliability

The human validation reviews were not conducted blind to pipeline condition, and inter-rater reliability (IRR) was not formally measured. Future work should incorporate blinded evaluation and IRR scoring for ambiguous (BORDERLINE) citations.

#### 4.8.13 Cost Figure Uncertainty

API costs for the VaaS pipeline are reported as *<*$1 per gene review, derived from downloaded Claude Sonnet API cost data. A precise total cannot be isolated from aggregate billing records because the AI fleet ran multiple projects in parallel during the production period. The *<*$1/gene figure is an upper bound; actual per-gene cost for VaaS-specific work is lower.

The Layer Contribution Summary table is reproduced below for reference:

**Table 20.** Pipeline layer contribution summary across VaaS conditions. ^*†*^AI self-assessed. ^*‡*^Intercepted, not in final output. **Note:** C1–C4 rows: prospective ablation data from VaaS-RIKER2 (*n* = 160 runs per condition). “Retrieval only” and “+L1 Aletheia” rows: reconstructed estimates from production logs. C2 rows: AI self-assessment, not live verification.

| Pipeline Configuration | Type I % | Type II % | Source |
| --- | --- | --- | --- |
| C1: No VaaS (unguided) | 1.4% | 95.9% | RIKER2, live-verified |
| C2: Corrections list only | 19.0%<sup>†</sup> | 15.1%<sup>†</sup> | RIKER2, AI self-assessed |
| C3: Verification only | 0.0% | 0.0% | RIKER2, live-verified |
| C4: Full VaaS protocol | 0.0% | 6.5%<sup>‡</sup> | RIKER2, live-verified |
| Retrieval only (no verification) | ~N/A | ~25% | Estimated from prod logs |
| +L1 Aletheia (existence check) | ~12% | ~12% | Estimated from prod logs |
| +PMID verification (live fetch + title) | ~2% | ~2% | Estimated from prod logs |

### 4.9 The Human-AI Partnership: Where the Loop Must Close

#### 4.9.1 Human Roles in the Pipeline

The corrections list grew because a human scientist (Dr. Stephen Ekker, Ph.D.) had domain knowledge sufficient to recognize when a stated drug approval sounded wrong, flagged it, and the catch was incorporated into the system. The self-improving loop is only as good as the human catches that seed it.

Domain expertise is not optional in the VaaS architecture. Human scientists contribute: (1) identification of novel error patterns (e.g., the first observation that Lumevoq’s EMA approval had been withdrawn); (2) verification of borderline citation decisions; (3) interpretation of multi-study findings; and (4) final sign-off on database entries used for healthcare support.

#### 4.9.2 Where AI Contribution Is Essential

##### Scale

No human team could read, synthesize, and correctly cite primary literature for 324 disease entries at less than $1 in API costs per detailed assessment — a total under $324 for the full database.

##### Consistency

AI agents apply the same verification protocol to every citation without fatigue, attention drift, or confirmation bias.

##### Citation coverage

The pipeline processes thousands of PMID candidates per gene batch, retaining only verified hits — a throughput impossible for manual review.

##### First-draft speed

A structured rare disease gene review — including inheritance, clinical features, therapeutic landscape, and zebrafish model data — is generated in approximately 8 minutes per entry.

#### 4.9.3 A Practical Model for Human-AI Scientific Partnership

## 5 Conclusions

A rigorous, multi-layer AI science quality pipeline can achieve near-zero citation fabrication rates and substantially reduce drug approval hallucinations at production scale. The VaaS-RIKER2 prospective benchmark provides the first controlled ablation study of such a pipeline in rare disease AI synthesis (757 total runs: 640 Claude arm + 117 open-weight model arm), confirming three of four prospectively defined hypotheses and establishing:

1. **Structural universality:** Wrong-topic citation hallucination is model-agnostic. C1 (Claude Sonnet) = 95.9% Type II; Buster OW arm = 81–87% Type II.
2. **Verification is the load-bearing gate:** C3 (live PMID verification only) achieves 0.0%/0.0% by catching wrong-topic and non-existent candidates before output.
3. **Corrections improve efficiency, not accuracy alone:** C2 reduces Type II from 95.9% to 15.1% but cannot achieve near-zero without verification. C4 achieves near-zero with higher citation yield than verification alone.
4. **Temperature invariance under full protocol:** C4 is completely temperature-invariant (H2 confirmed). Temperature matters for corrections-only conditions (H1 partial); it is irrelevant under full VaaS.

**Table 21.** Division of labor in the VaaS human-AI scientific partnership.

| Task | AI Role | Human Role |
| --- | --- | --- |
| Literature mining | Exhaustive PubMed candidate retrieval | Define search scope and relevance criteria |
| Citation verification | Live fetch + topic check for every PMID | Review borderline cases |
| Drug approval status | Parametric recall + corrections list | Identify novel errors, update corrections list |
| Error pattern recognition | Flag anomalies via First Law trigger | Interpret patterns, add to corrections list |
| Clinical interpretation | Draft synthesis from verified papers | Final clinical accuracy review |
| Domain knowledge | Apply prior corrections | Source and validate domain knowledge |

The key architectural elements validated across production and prospective benchmarks are:

- **The First Law:** Scientific integrity as an identity-level constraint, not a task instruction.
- **Live verification:** Direct HTTP fetch for every PMID, not parametric recall.
- **Self-improving corrections injection:** A living document of domain-specific known failures.
- **Subagent isolation:** Fresh context per entry eliminates cross-contamination.
- **Cross-model validation:** Training corpus diversity as an ensemble advantage.
- **Tiered risk assignment:** Proportional scrutiny for proportional stakes.
- **Multi-layer manifest QC:** Catches duplicate synonyms and nonexistent genes before any review is written.
- **Hallucination topology awareness:** Tier-stratified protocol design addressing qualitatively distinct failure modes (Tier 1: paralog confusion; Tier 3: wholesale fabrication).
- **Topic verification protocol:** Explicit YES/NO/BORDERLINE classification of every candidate citation against the target gene (Section 2.4).
- **Prospectively defined benchmarking:** Validation against held-out gene sets with temperature sweeps and open-weight model comparison.

These results demonstrate that LLM citation hallucination is a structural, model-agnostic phenomenon that the VaaS pipeline consistently reduces to near zero. The pipeline is deployable at production scale for less than $1 per gene review in API costs. The human-AI partnership remains essential: the pipeline amplifies human domain expertise rather than replacing it.

**Future directions** include cross-architecture comparison with GPT-4o and Gemini, prospective expansion of the batch pipeline to additional gene families, independent re-scoring of the OW arm with the full VaaS verification protocol, and a formal examination of the pipeline’s performance on the broader nuclear-encoded mitochondrial disease gene space. (For context, a comprehensive nuclear-encoded mitochondrial disease gene list (Stenton and Prokisch, 2020) provides one candidate universe for future expansion; that gene list is not directly analyzed in the current paper.)

## Data Availability

All data produced in the present study are available upon reasonable request to the authors.

## Author Contributions

Co-first authorship (†): AS and MP contributed equally.

**Stephen C. Ekker** — Collaborative, Domain Validation, Novel Error Pattern Identification, Writing — Review & Editing, Corresponding Author.

**Ankit Sabharwal** — Conceptualization (Rare Disease Database), Data Curation, Formal Analysis, Investigation, Methodology, Writing — Original Draft (RDD companion paper), Writing — Review & Editing.

**Anna Carrano** — Human Validation (gene assessment cohort), Writing — Review & Editing.

**Maarten Rotman** — Human Validation (gene assessment cohort), Writing — Review & Editing.

**Milit Patel** — Worked with SCE to establish the first AI Scientist for initial training, Writing — Review & Editing.

**Wesley Wierson** — Investigation, Methodology, Writing — Review & Editing.

## Acknowledgments

Uvy (UViiVe AI Scientist) and Atlas (UViiVe AI Scientist) contributed 74 of 225 database entries under Wave 2 protocols. Automated pipeline agents (Pip, Jimmy, Atlas) contributed batch validation data for the v2.0 expansion pipeline, including the QC catches (ADCK3 duplicate detection, BCS1L2 nonexistent gene detection, DMAC1 fabricated PMID/OMIM# detection) documented in Section 3.9. Atlas contributed open-weight model simulation data and conducted the independent L3 citation audit of Wave 3 (March 15, 2026). Buster (UViiVe GPU node, NVIDIA RTX 3080 Ti) contributed the formal VaaS-RIKER2 open-weight arm runs on dedicated hardware (Ollama inference; `llama3.2`, `qwen2.5:14b`, `mistral:7b`). All fleet agents operated under identical quality standards and the same epistemic integrity constraint.

## AI Contribution Statement

This manuscript was produced through a structured human-AI collaboration (UViiVe / UT Austin), using UViiVe’s multi-agent orchestration framework. The following AI agents made substantive, documented contributions:

**Zevo** (Center for Rare Disease deployed AI Lead Scientist; Claude Sonnet 4 backbone, Amazon Bedrock) — Primary manuscript author across drafts v1–v21. Performed literature retrieval, citation verification, data analysis, figure legend drafting, and manuscript text generation. Designed and executed Wave 2 parallel production. Led VaaS-RIKER2 prospective benchmark design and execution.

**Atlas** (UViiVe AI Scientist; Claude Sonnet 4 backbone, Amazon Bedrock; Mac Studio M3 Ultra, 192GB RAM) — Contributed 74 Wave 2 gene review entries (cardiac, immune, and renal disease genes). Conducted the independent L3 citation audit of Wave 3 (100 genes, 179 PMIDs; March 15, 2026). Contributed open-weight model simulation data.

**Uvy** (UViiVe AI Scientist; Claude Sonnet 4 backbone, Amazon Bedrock) — Contributed 74 Wave 2 gene review entries (lysosomal, neurological, and sensory disease genes).

**Buster** (UViiVe Protein Design Engineer; Claude Sonnet 4 backbone, Amazon Bedrock; Framework laptop, NVIDIA RTX 3080 Ti 12GB VRAM, Ollama inference) — Executed the formal VaaS-RIKER2 open-weight arm: 117 runs across three model architectures at four temperature settings. All inference conducted on-device with no internet access during generation. Identified and documented the CHCHD10 scoring anomaly.

**Pip** (UViiVe test agent; Claude Sonnet 4 backbone, Amazon Bedrock) — Contributed batch validation pipeline data including three automated QC catches. Executed MedHallu Run 3 (independent cold-SOP replication, *N* = 100 hard-tier items, Claude Sonnet 4.5), achieving 100% accuracy.

**Jimmy** (UViiVe coordination agent; Claude Sonnet 4 backbone, Amazon Bedrock) — Contributed batch validation pipeline data. Participated in automated quality gate operations during production scaling.

All fleet agents operated under identical quality standards and the same epistemic integrity constraint (“The First Law”). All AI-generated text was reviewed and approved by Dr. Ekker prior to submission.

**Tool/model information:** Primary generation — Claude Sonnet 4 (Anthropic, via Amazon Bedrock, us-east-1); open-weight model arm — `llama3.2:3b`, `qwen2.5:14b`, `mistral:7b` (Ollama, NVIDIA RTX 3080 Ti, Buster node); cross-validation — Kimi K2.5 (Moonshot AI). Pipeline orchestration — OpenClaw (v2026.2.24+).

## Supplementary Material

### Box S3: Full Topic Verification Prompt

*Purpose:* Applied to every PMID candidate after live-fetch existence confirmation. Determines whether a verified paper directly studies the target gene.

**Verbatim prompt:** “Given the following paper abstract, does this paper directly study `[GENE_SYMBOL]`? Answer YES, NO, or BORDERLINE with one sentence of explanation. Abstract: `[ABSTRACT]`”

### Box S4: Cross-Validation Prompt

*Purpose:* Applied when two independent gene review summaries are available for the same gene. Identifies claims present in one summary but absent or contradicted in the other.

**Verbatim prompt:** “Given two independent summaries of gene `[GENE]` disease associations, identify any claims present in one summary but absent or contradicted in the other. For each discrepancy, classify it as: (A) factual contradiction; (B) coverage gap; or (C) confidence difference. For each identified discrepancy, state which summary you consider more likely to be accurate and why. Gene: `[GENE]`. Summary 1: `[SUMMARY_1]`. Summary 2: `[SUMMARY_2]`.”

### Box S5: VaaS-RIKER2 Scoring Protocol

**Type I error** (`hall_rate_type1`): Fraction of cited PMIDs confirmed non-existent by live PubMed fetch. Counted at the run level; averaged across runs for aggregate rates.

**Type II error** (`wrong_topic_rate_type2`): Fraction of cited PMIDs confirmed to exist but scoring NO on the topic verification prompt (Section 2.4). Counted at the run level.

**C2 special case:** No live fetch performed in C2. Type I represents model self-reported calibration uncertainty. Type II represents model-assessed wrong-topic likelihood. These figures are not directly comparable to C1/C3/C4 live-verified rates.

**Ground-truth (GT) recall:** Fraction of manifest-verified PMIDs (`manifest.json`, 5 per gene) recovered in the cited output. Low GT recall (3% in C3/C4) reflects that independent parametric citation generation does not systematically produce the same specific PMIDs as manual curation — a separate issue from hallucination rate.

## References

Mehul Bhattacharyya, Valerie M. Miller, Debjani Bhattacharyya, and Larry E. Miller. High Rates of Fabricated and Inaccurate References in ChatGPT-Generated Medical Content. Cureus, 15(5):e39238, May 2023. ISSN 2168-8184. doi: 10.7759/cureus.39238.

Moses Boudourides. Structural Hallucination in Large Language Models: A Network-Based Evaluation of Knowledge Organization and Citation Integrity, 2026. https://arxiv.org/abs/2603.01341. Version Number: 1.

Ishaan Gangwani and Aayam Bansal. CiteGuard: Retrieval-augmented citation verification for LLM-powered peer review. In Workshop: AI for Science, NeurIPS 2025 (Spotlight), 2025. Proceedings ID 125912, https://neurips.cc/virtual/2025/125912.

Sharon Goldman. NeurIPS, one of the world’s top academic AI conferences, accepted research papers with 100+ AI-hallucinated citations, new report claims. Fortune, January 2026. https://fortune.com/2026/01/21/neurips-ai-conferences-research-papers-hallucinations/. News report.

Ziwei Ji, Nayeon Lee, Rita Frieske, Tiezheng Yu, Dan Su, Yan Xu, Etsuko Ishii, Ye Jin Bang, Andrea Madotto, and Pascale Fung. Survey of Hallucination in Natural Language Generation. ACM Computing Surveys, 55(12):1–38, December 2023. ISSN 0360-0300, 1557-7341. doi: 10.1145/3571730. https://dl.acm.org/doi/10.1145/3571730.

Sota Nishisako, Takahiro Higashi, and Fumihiko Wakao. Reducing Hallucinations and Trade-Offs in Responses in Generative AI Chatbots for Cancer Information: Development and Evaluation Study. JMIR Cancer, 11:e70176, September 2025. ISSN 2369-1999. doi: 10.2196/70176.

OpenAI, Josh Achiam, Steven Adler, Sandhini Agarwal, Lama Ahmad, Ilge Akkaya, Florencia Leoni Aleman, Diogo Almeida, Janko Altenschmidt, Sam Altman, Shyamal Anadkat, Red Avila, Igor Babuschkin, Suchir Balaji, Valerie Balcom, Paul Baltescu, Haiming Bao, Mohammad Bavarian, Jeff Belgum, Irwan Bello, Jake Berdine, Gabriel Bernadett-Shapiro, Christopher Berner, Lenny Bogdonoff, Oleg Boiko, Madelaine Boyd, Anna-Luisa Brakman, Greg Brockman, Tim Brooks, Miles Brundage, Kevin Button, Trevor Cai, Rosie Campbell, Andrew Cann, Brittany Carey, Chelsea Carlson, Rory Carmichael, Brooke Chan, Che Chang, Fotis Chantzis, Derek Chen, Sully Chen, Ruby Chen, Jason Chen, Mark Chen, Ben Chess, Chester Cho, Casey Chu, Hyung Won Chung, Dave Cummings, Jeremiah Currier, Yunxing Dai, Cory Decareaux, Thomas Degry, Noah Deutsch, Damien Deville, Arka Dhar, David Dohan, Steve Dowling, Sheila Dunning, Adrien Ecoffet, Atty Eleti, Tyna Eloundou, David Farhi, Liam Fedus, Niko Felix, Simón Posada Fishman, Juston Forte, Isabella Fulford, Leo Gao, Elie Georges, Christian Gibson, Vik Goel, Tarun Gogineni, Gabriel Goh, Rapha Gontijo-Lopes, Jonathan Gordon, Morgan Grafstein, Scott Gray, Ryan Greene, Joshua Gross, Shixiang Shane Gu, Yufei Guo, Chris Hallacy, Jesse Han, Jeff Harris, Yuchen He, Mike Heaton, Johannes Heidecke, Chris Hesse, Alan Hickey, Wade Hickey, Peter Hoeschele, Brandon Houghton, Kenny Hsu, Shengli Hu, Xin Hu, Joost Huizinga, Shantanu Jain, Shawn Jain, Joanne Jang, Angela Jiang, Roger Jiang, Haozhun Jin, Denny Jin, Shino Jomoto, Billie Jonn, Heewoo Jun, Tomer Kaftan, Łukasz Kaiser, Ali Kamali, Ingmar Kanitscheider, Nitish Shirish Keskar, Tabarak Khan, Logan Kilpatrick, Jong Wook Kim, Christina Kim, Yongjik Kim, Jan Hendrik Kirchner, Jamie Kiros, Matt Knight, Daniel Kokotajlo, Łukasz Kondraciuk, Andrew Kondrich, Aris Konstantinidis, Kyle Kosic, Gretchen Krueger, Vishal Kuo, Michael Lampe, Ikai Lan, Teddy Lee, Jan Leike, Jade Leung, Daniel Levy, Chak Ming Li, Rachel Lim, Molly Lin, Stephanie Lin, Mateusz Litwin, Theresa Lopez, Ryan Lowe, Patricia Lue, Anna Makanju, Kim Malfacini, Sam Manning, Todor Markov, Yaniv Markovski, Bianca Martin, Katie Mayer, Andrew Mayne, Bob McGrew, Scott Mayer McKinney, Christine McLeavey, Paul McMillan, Jake McNeil, David Medina, Aalok Mehta, Jacob Menick, Luke Metz, Andrey Mishchenko, Pamela Mishkin, Vinnie Monaco, Evan Morikawa, Daniel Mossing, Tong Mu, Mira Murati, Oleg Murk, David Mély, Ashvin Nair, Reiichiro Nakano, Rajeev Nayak, Arvind Neelakantan, Richard Ngo, Hyeonwoo Noh, Long Ouyang, Cullen O’Keefe, Jakub Pachocki, Alex Paino, Joe Palermo, Ashley Pantuliano, Giambattista Parascandolo, Joel Parish, Emy Parparita, Alex Passos, Mikhail Pavlov, Andrew Peng, Adam Perelman, Filipe de Avila Belbute Peres, Michael Petrov, Henrique Ponde de Oliveira Pinto, Michael, Pokorny, Michelle Pokrass, Vitchyr H. Pong, Tolly Powell, Alethea Power, Boris Power, Elizabeth Proehl, Raul Puri, Alec Radford, Jack Rae, Aditya Ramesh, Cameron Raymond, Francis Real, Kendra Rimbach, Carl Ross, Bob Rotsted, Henri Roussez, Nick Ryder, Mario Saltarelli, Ted Sanders, Shibani Santurkar, Girish Sastry, Heather Schmidt, David Schnurr, John Schulman, Daniel Selsam, Kyla Sheppard, Toki Sherbakov, Jessica Shieh, Sarah Shoker, Pranav Shyam, Szymon Sidor, Eric Sigler, Maddie Simens, Jordan Sitkin, Katarina Slama, Ian Sohl, Benjamin Sokolowsky, Yang Song, Natalie Staudacher, Felipe Petroski Such, Natalie Summers, Ilya Sutskever, Jie Tang, Nikolas Tezak, Madeleine B. Thompson, Phil Tillet, Amin Tootoonchian, Elizabeth Tseng, Preston Tuggle, Nick Turley, Jerry Tworek, Juan Felipe Cerón Uribe, Andrea Vallone, Arun Vijayvergiya, Chelsea Voss, Carroll Wainwright, Justin Jay Wang, Alvin Wang, Ben Wang, Jonathan Ward, Jason Wei, CJ Weinmann, Akila Welihinda, Peter Welinder, Jiayi Weng, Lilian Weng, Matt Wiethoff, Dave Willner, Clemens Winter, Samuel Wolrich, Hannah Wong, Lauren Workman, Sherwin Wu, Jeff Wu, Michael Wu, Kai Xiao, Tao Xu, Sarah Yoo, Kevin Yu, Qiming Yuan, Wojciech Zaremba, Rowan Zellers, Chong Zhang, Marvin Zhang, Shengjia Zhao, Tianhao Zheng, Juntang Zhuang, William Zhuk, and Barret Zoph. GPT-4 Technical Report, 2023. https://arxiv.org/abs/2303.08774. Version Number: 6.

Shrey Pandit, Jiawei Xu, Junyuan Hong, Zhangyang Wang, Tianlong Chen, Kaidi Xu, and Ying Ding. MedHallu: A Comprehensive Benchmark for Detecting Medical Hallucinations in Large Language Models, 2025. https://arxiv.org/abs/2502.14302. Version Number: 1.

Haosheng Qian, Yixing Fan, Jiafeng Guo, Ruqing Zhang, Qi Chen, Dawei Yin, and Xueqi Cheng. VeriCite: Towards Reliable Citations in Retrieval-Augmented Generation via Rigorous Verification, 2025. https://arxiv.org/abs/2510.11394. Version Number: 1.

JV Roig. How Much Do LLMs Hallucinate in Document Q&A Scenarios? A 172-Billion-Token Study Across Temperatures, Context Lengths, and Hardware Platforms, 2026. https://arxiv.org/abs/2603.08274. Version Number: 1.

Shirin Salehi, Yash Singh, Kimberly K. Horst, Quincy A. Hathaway, and Bradley J. Erickson. Agentic AI and large language models in radiology: Opportunities and hallucination challenges. Bioengineering, 12(12):1303, 2025. doi: 10.3390/bioengineering12121303.

Sarah L. Stenton and Holger Prokisch. Genetics of mitochondrial diseases: Identifying mutations to help diagnosis. EBioMedicine, 56:102784, 2020. doi: 10.1016/j.ebiom.2020.102784.

